# Early warning signals predict emergence of COVID-19 waves

**DOI:** 10.1101/2021.06.24.21259444

**Authors:** Duncan A. O’Brien, Christopher F. Clements

## Abstract

Early warning signals (EWSs) aim to predict changes in complex systems from phenomenological signals in time series data. These signals have recently been shown to precede the initial emergence of disease outbreaks, offering hope that policy makers can make predictive rather than reactive management decisions. Here, using daily COVID-19 case data in combination with a novel, sequential analysis, we show that composite EWSs consisting of variance, autocorrelation, and return rate not only pre-empt the initial emergence of COVID-19 in the UK by 14 to 29 days, but also the following wave six months later. We also predict there is a high likelihood of a third wave as of the data available on 9th June 2021. Our work suggests that in highly monitored disease time series such as COVID-19, EWSs offer the opportunity for policy makers to improve the accuracy of time critical decisions based solely upon surveillance data.

## Introduction

As with many natural systems, the emergence of infectious disease is often sudden and non-linear, making it difficult for policy makers to identify and appropriately manage threats [1,2]. Balancing the health risks posed by these novel diseases and the possible economic impact of imposing mitigation strategies is therefore a complicated process ultimately dependent on the timing of strategy implementation [3].

The severe acute respiratory syndrome-coronavirus 2 (SARS-CoV-2), which causes the Coronavirus disease 2019 (COVID-19), exemplifies this challenge. There has been widespread criticism, in response to the harm the pandemic has inflicted, of the speed and severity of the national strategies to the virus threat [4,5]. To date, governments have imposed a spectrum of clinical (e.g. intensive care unit construction, quarantine), non-pharmaceutical (social distancing, large-scale lockdowns etc.) and, most recently, vaccination-based strategies, with the timing of implementation dramatically influencing the case curve between and within legislative regions [6]. Non-pharmaceutical interventions (NPIs) represent the severest but highly effective epidemic controls, with previous analysis suggesting their implementation stabilised European reproduction number below one, and reduced the number of deaths compared to scenarios with no intervention [7]. The optimum moment of action is unclear however, with suggestions that, due to periods of cryptic transmission [8,9], strong NPIs two weeks earlier than performed may have halved cumulative deaths [10,11]. Identifying this cryptic window would therefore improve strategy decisions both in enforcing initial NPIs in response to the virus’ emergence and possible future NPIs based upon the relevant COVID-19 case situation.

Unfortunately, the causes of disease emergence and/or re-emergence often appear idiosyncratic [12], requiring the use of context-specific models [13,14]. These models are powerful tools and have become keystones during the COVID-19 pandemic response, but are restricted by data availability [15,16], potential for lack of transparency [17] and our mechanistic understanding of the system [18]. Due to these difficulties, there have been alternative suggestions to consider disease emergence as critical transitions [19] where a forcing pressure, such as host movement or pathogen evolution, drive the system towards a threshold. If emergence is considered in this manner, then a suite of alternative methods based upon the concept of critical slowing down (CSD) become applicable in the identification of transitions in disease and COVID-19 dynamics.

Critical slowing down represents the compromised ability of a system to recover from perturbation as it approaches a threshold, at which a small perturbation in the state triggers a positive feedback loop and a system shift [20]. From this phenomena, various mechanism-independent and summary statistic based indicators have been identified across a range simulated [21,22] and empirical [23,24] studies. In disease systems specifically, CSD was established as tracking a transcritical transition in the effective reproductive number, *R*_eff_, [25] or number of secondary infections an infectious individual causes. Below one, secondary infection is unlikely whereas above one, transmission is common and an outbreak of sustained disease occurs. The period where *R*_eff_ increases towards one is represented by a region of ‘stuttering’/cryptic transmission [26] during which, CSD also increases [25,27]. The ‘Early Warning Signal’ (EWS) summary statistics based upon CSD will therefore precede the rapid increase in cases at the beginning of a disease outbreak; before *R*_eff_ exceeds a value of one [28].

Early warning signals have been shown to predict the emergence of diseases in both empirical and simulation studies [25,29,30]. Variance, autocorrelation at lag1, and decay time all increased prior to malaria resurgence in Kenya [29] and before the initial emergence of COVID-19 in seven of the nine countries assessed [31]. However, a critical use of EWSs is not only to predict initial emergence, but also successive re-emergences, with no method currently developed to achieve this. Thus, COVID-19 represents a unique opportunity to test the efficacy of EWSs to predict multiple sequential outbreaks by assessing the wave-like dynamics expressed in most countries [32].

Here we test whether CSD based EWSs can predict sequential transitions in COVID-19 daily case numbers, developing a novel methodology to detect and subset data into successive waves of infection. Using the suggested framework, we show evidence that EWS can be identified prior to each of the two previous COVID-19 waves experienced by the United Kingdom and predicts a third as of the data available on 9^th^ June 2021. Our results provide suggestions on how to use EWSs in a management scenario, where decisions must be made as data are collected, rather than post hoc.

## Methods

### Data Availability

Daily COVID-19 case data was collected from the UK government’s coronavirus data portal (https://coronavirus.data.gov.uk) and World Health Organisation’s dashboard (https://covid19.who.int/info/), spanning 30^th^ January 2020 to 9^th^ June 2021. Uniquely, we analysed positive, daily COVID-19 cases rather than cumulative cases as performed by other studies [31], thus allowing us to attempt the prediction of sequential COVID-19 outbreaks. Case data has previously been criticised for its inaccuracies resulting from incomplete testing and cryptic cases [7], but we wanted to explore the viability of EWSs using the most universally collected and interpreted data type. The data were consequently analysed in its raw form with no detrending performed.

### COVID-19 Wave Identification

To assess the instantaneous increase and decline of COVID-19 cases and define ‘waves’, generalised additive mixed effect models (GAMMs) were fitted to daily cases using the R language [33] package ‘mgcv’ [34], following the method employed by [35]. The optimum number of spline knots was selected via generalized cross-validation [34] with a supplementary ARIMA correlation structure by time step fitted to account for the autocorrelation inherent in time series. A day-of-the-week smooth was also fitted to account for potential variation in testing rates over the weekend. The time series’ inflection points, indicating the onset and recovery of COVID-19 waves, were defined by the significant differing of the fitted model’s first derivatives from zero, as assessed by 95% pointwise confidence intervals. A wave onset was identified by a significant, positive first derivative maintained for seven data points after a period of stationarity, whereas the subsidence of the wave was identified by a stationary period following a succession of significant, negative derivatives. The choice of a seven datapoint persistence was informed by the upper incubation period of the SARS-CoV-2 virus (μ = 5.8 days; 95% CI: 5.0 to 6.7) [36]. First derivatives were extracted from the GAMM fit using the ‘derivatives’ function in the ‘gratia’ package [37] with the direction of derivative change performed by a custom function published by [35].

### Early Warning Signal Calculation

The presence of EWSs was then calculated using the framework suggested by [38] and developed in [24,39]. Briefly, this approach differs from other EWS methodologies [40] by assessing time series change via a composite metric consisting of multiple indicators. Here, we focussed on the two most commonly used EWS indicators, variance (represented by the standard deviation, SD) and autocorrelation at first lag (acf) [41], as well as the return rate (rr), which represents the inherent quantity of interest in indicators based upon CSD [42]. Each indicator was normalised by subtracting its expanding mean from its calculated value at time *t* before dividing by its expanding standard deviation [24]. A composite metric was then constructed by summing all individual indicator values calculated per time point. An EWS was therefore considered present when the composite metric exceeded its expanding mean by 2σ [38]. The 2σ threshold was chosen based upon its equivalency to a 95% confidence interval and repeatedly favourable performance compared to other threshold levels [24,39].

As the expanding mean of the indicator is the basis of assessment, a previous wave will often mask the appearance of second (Supplementary Figure 1). Consequently, once a wave subsided, as assessed by GAMM first derivatives, the data were cut and the EWS assessment restarted, truncating the time series from the point of wave end. This resulted in a series of EWS assessments, each for a specific wave and independent from previous waves. Similarly, the expanding window approach is susceptible to false positive signals at the start of assessment due to the short time series length and high variability when few data points are supplied to the algorithm. To mitigate this, a seven time step burn-in period was introduced (the upper duration of COVID-19 incubation) before metric strength was assessed to ‘train’ the signals.

To compare the robustness of indicators, a ‘warning’ was considered present under two scenarios. First, when the initial signal was identified, and second (to represent a conservative assessment) whether signals were detected for seven consecutive time steps. Evidence suggests that a persistent signal for two time steps is sufficient to reduce the frequency of false positives [43], but as that work focussed on a systems’ recovery, a stricter persistence was assessed here. From the calculated indicators, we present both the individual indicator strengths over time as well as the difference between the time-of-first-detection and the onset of each wave estimated from the GAMM derivatives. We also identify the superior indicator or combination of indicators for specifically pre-empting COVID-19 waves.

## Results

Generalised additive mixed effect models predicted two significantly increasing regions in daily UK COVID-19 cases. These regions correspond to the onset of Waves 1 and 2 beginning on 17^th^ March 2020 and 9^th^ September 2020 respectively. From this prediction, two restarts of the EWS analysis were performed on 18^th^ June 2020 and 1st April 2021 following the subsidence of each wave.

All early warning signal (EWS) indicators, excluding return rate (rr), increased and exceeded the 2σ threshold at least once prior to the appearance of COVID-19 Waves 1 and 2 (Figure 1, Supplementary Table 1a). Time-of-first-detection was consistently two weeks or earlier than the predicted wave onset, with Wave 2 being pre-empted earlier than Wave 1 (Supplementary Table 1). No third wave was strongly identified by the GAMM first derivatives but the majority of indicators indicate the potential for an oncoming third wave (Figure 2a).

**Figure 1.**
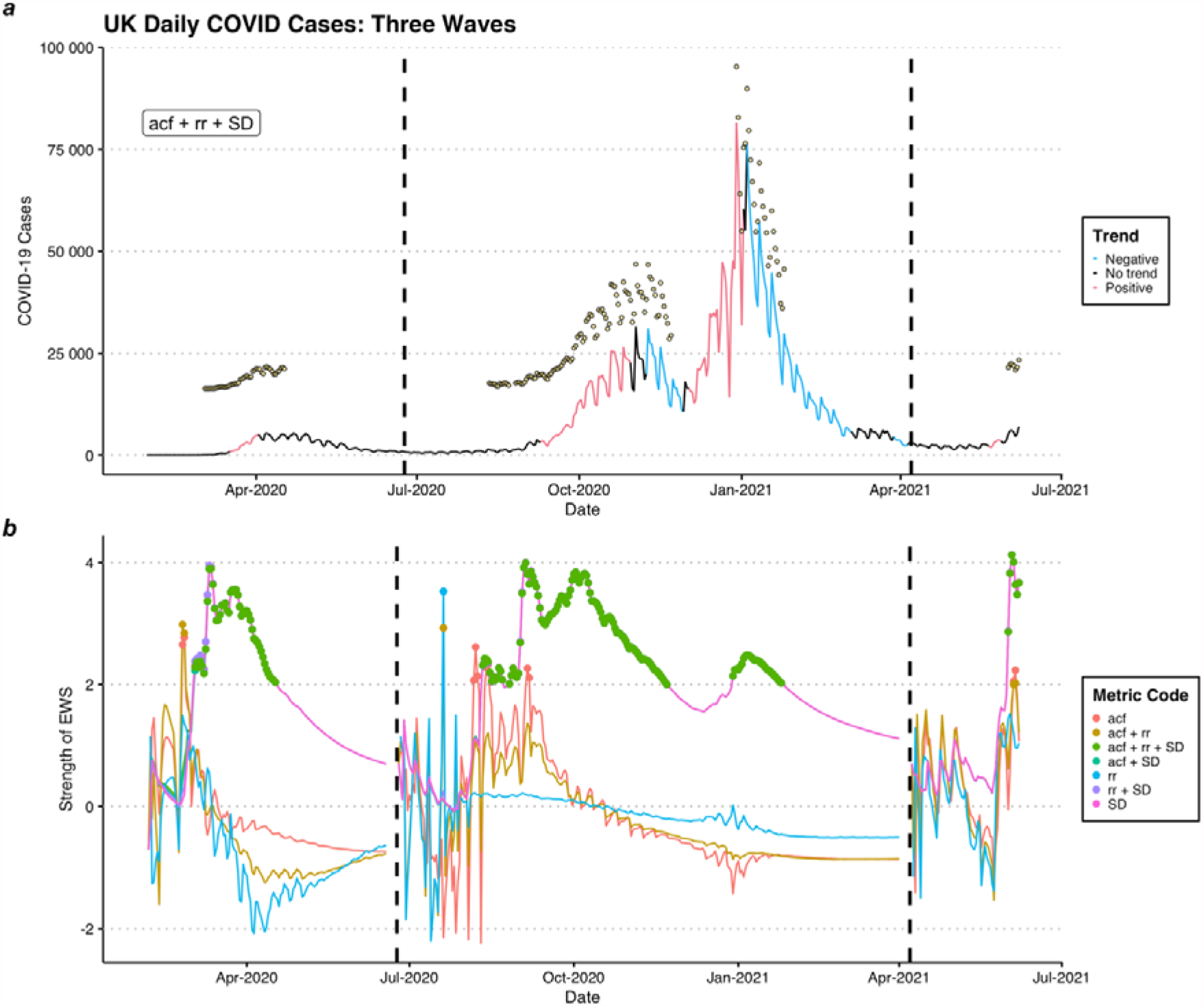
Time series of a) the daily UK COVD-19 cases and GAMM predicted trends, where yellow points represent detected early warning signals (EWSs) from the triple composite indicator ‘acf + rr + SD’, and b) individual EWS indicator strengths, where coloured dots indicate time points exceeding the 2σ threshold. Dashed, vertical lines indicate the initialisation of EWS reassessment.

**Figure 2.**
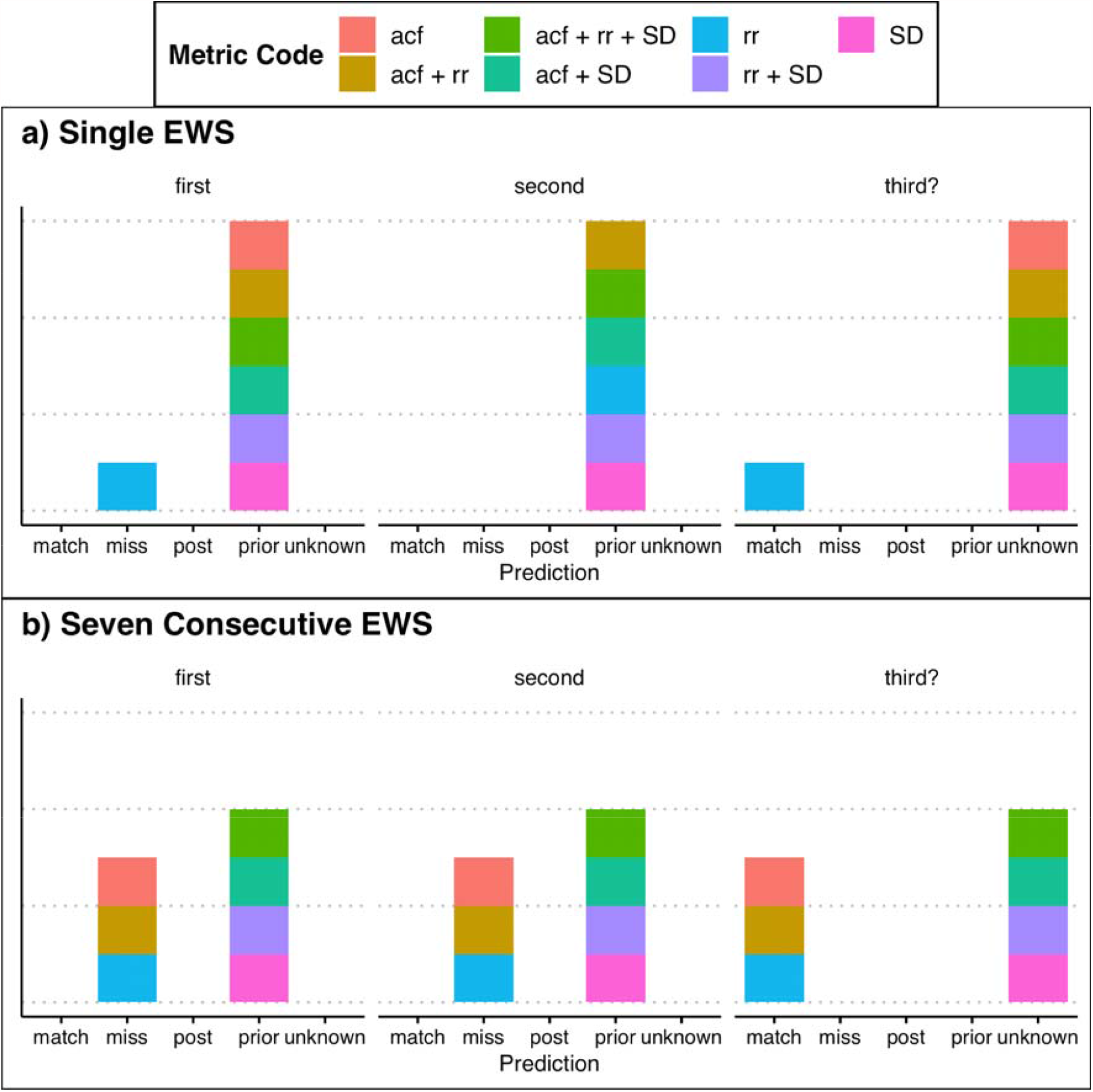
Predictions of individual early warning signal indicators based upon the difference between their time-of-first-detection and estimated time-of-wave-onset under a) a lenient (single signal) and b) a strict definition of a warning (seven consecutive signals). ‘*Match*’ represents identical time-of-first-detection and time-of-wave-onset, ‘*miss*’ being no warning despite the presence of a wave, ‘*post*’ is the identification of a warning after the time-of-wave-onset, ‘*prior*’ is the identification of a warning before the time-of-wave-onset and ‘*unknown*’ representing the identification of a warning with no apparent wave (i.e. potential for an oncoming wave). Indicators include: autocorrelation (acf), return rate (rr) and variance (SD).

However, when a warning is only accepted as a sequence of seven consecutive signals, many of the indicators are no longer interpretable as warnings. For this more conservative assessment, indicators incorporating variance (SD) become the sole signal providers (Figure 2b, Supplementary Table 1b) and resultingly, all times-of-first-detection were identical. Wave 2 remains better pre-empted than Wave 1 for successful indicators, as does the possibility of a third wave.

Similar results for international COVID-19 cases were observed (Supplementary Figure 2) with variance remaining the most robust predictor of both emergence and re-emergence. Successive waves remain well predicted in many countries although in nations where daily variation is high and the time between waves is short, EWSs become delayed and do not pre-empt the wave onset.

## Discussion

These results show that Early Warning Signals (EWSs) based on the theory of critical slowing down (CSD) are sufficient to predict the emergence, and subsequent re-emergence, of COVID-19 in the UK and abroad. This is strengthened when time series are subset into individual waves, and the timing of subsets unbiasedly estimated using a GAMM approach. As all indicators increased prior to the onset of COVID-19 waves, there is agreement with the prediction that CSD is observable in the disease’s dynamics [25] and so represents a viable tool for policy makers to inform timely decisions.

With variance responding consistently under the strictest definition of a warning, this implies that the indicator is the most reliable for pre-empting disease. This is congruent with previous disease [29,44] and ecological [24,41] findings. The weakness of autocorrelation was unexpected considering those studies also identified autocorrelation as a reliable indicator, yet the robustness of the triple composite EWS regardless highlights the benefit of the composite metric approach developed by [38] over individual indicators. The degree of pre-emption reported here is particularly important as, statistically, the lag between transition and disease emergence is not fully understood [28] and the supposition that earlier non-pharmaceutical interventions (NPIs) may have dramatically improved death rates early in the pandemic [10]. We therefore suggest that composite EWSs are detectable sufficiently prior to COVID-19 waves to be suitable in the current monitoring toolbox for this and potential future pandemics.

Although this study provides evidence for the usefulness of EWS indicators, their application is often hampered by the quality of data available [45]. Epidemiological data can be particularly problematic due to the reporting style of practitioners. Case data can be aggregated in to weekly or monthly counts with the exact date of infection/recovery of an individual unknown, or, during the emergence of disease specifically, many cases will be cryptic due to a lack of testing or symptoms [7]. Whilst EWSs based upon disease incidence often display unique behaviour compared to the same indicators based upon disease prevalence, this study suggests EWSs can still detect critical transitions in daily data via an expanding window and is supported by other studies implementing the alternative rolling window approach [30,46]. COVID-19 case data from the UK does provide a best-case example for EWS assessment however, due to the frequency of testing and defined periods of stationarity enforced by NPIs, but as many governments alter their approach towards future pandemics [47], the quality of data may become universal. Even with contemporary levels of reporting variation between countries, EWSs do consistently detect the onset of waves (Supplementary Figure 2), though the strength of pre-emption varies.

In conclusion, the results of this study support the use of composite CSD indicators during COVID-19 monitoring, and likely other diseases with re-emergence dynamics. The two week plus pre-emption of wave onset is encouraging for their informing of policy, namely as a ‘preliminary analysis’ indicative of a system at risk and requiring intervention consideration. The detection of EWSs could thus be followed by efforts to specifically identify the underlying variables of the system that are changing. Although we advocate the use of sequential EWS assessment for disease monitoring, it is harder to apply this approach in multi-dimensional systems where a stationary period is likely unidentifiable. Ecosystems, for example, are constantly fluctuating in response to intrinsic or extrinsic drivers [48], with it unclear the minimum length of time series to definitively identify the system’s trend [49]. Nonetheless, if periods of stationarity can be identified, we believe sequential assessment is necessary for EWS usage and prevention of bias from historic transitions.

## Data Availability

Data was collected from the UK governments coronavirus data portal (https://coronavirus.data.gov.uk) and World Health Organisations dashboard (https://covid19.who.int/info/).

## Acknowledgements

We are thankful to the GW4+ FRESH Centre for Doctoral Training in Freshwater Biosciences and Sustainability for their support of this project.

## References

1. Howard CR, Fletcher NF. 2012 Emerging virus diseases: can we ever expect the unexpected? Emerg. Microbes Infect. 1, e46–e46. (doi:10.1038/emi.2012.47)

2. Morens DM, Folkers GK, Fauci AS. 2004 The challenge of emerging and re-emerging infectious diseases. Nature 430, 242–249. (doi:10.1038/nature02759)

3. Anderson RM, Heesterbeek H, Klinkenberg D, Hollingsworth TD. 2020 How will country-based mitigation measures influence the course of the COVID-19 epidemic? Lancet 395, 931–934. (doi:10.1016/S0140-6736(20)30567-5)

4. Cairney P. 2021 The UK government’s COVID-19 policy: assessing evidence-informed policy analysis in real time. Br. Polit. 16, 90–116. (doi:10.1057/s41293-020-00150-8)

5. Devine D, Gaskell J, Jennings W, Stoker G. 2020 Trust and the coronavirus pandemic: What are the consequences of and for trust? An early review of the literature. Polit. Stud. Rev. 19, 274–285. (doi:10.1177/1478929920948684)

6. Prem K et al. 2020 The effect of control strategies to reduce social mixing on outcomes of the COVID-19 epidemic in Wuhan, China: a modelling study. Lancet Public Heal. 5, e261–e270. (doi:10.1016/S2468-2667(20)30073-6)

7. Flaxman S et al. 2020 Estimating the effects of non-pharmaceutical interventions on COVID-19 in Europe. Nature 584, 257–261. (doi:10.1038/s41586-020-2405-7)

8. Bedford T et al. 2020 Cryptic transmission of SARS-CoV-2 in Washington state. Science (80-.). 370, 571 LP –575. (doi:10.1126/science.abc0523)

9. Li R, Pei S, Chen B, Song Y, Zhang T, Yang W, Shaman J. 2020 Substantial undocumented infection facilitates the rapid dissemination of novel coronavirus (SARS-CoV-2). Science (80-.). 368, 489 LP –493. (doi:10.1126/science.abb3221)

10. Ragonnet-Cronin M et al. 2021 Genetic evidence for the association between COVID-19 epidemic severity and timing of non-pharmaceutical interventions. Nat. Commun. 12, 2188. (doi:10.1038/s41467-021-22366-y)

11. Hale T, Hale AJ, Kira B, Petherick A, Phillips T, Sridhar D, Thompson RN, Webster S, Angrist N. 2020 Global assessment of the relationship between government response measures and COVID-19 deaths. medRxiv, 2020.07.04.20145334. (doi:10.1101/2020.07.04.20145334)

12. Jones KE, Patel NG, Levy MA, Storeygard A, Balk D, Gittleman JL, Daszak P. 2008 Global trends in emerging infectious diseases. Nature 451, 990–993. (doi:10.1038/nature06536)

13. Kim Y, Lee S, Chu C, Choe S, Hong S, Shin Y. 2016 The characteristics of Middle Eastern Respiratory Syndrome coronavirus transmission dynamics in South Korea. Osong Public Heal. Res. Perspect. 7, 49–55. (doi:https://doi.org/10.1016/j.phrp.2016.01.001)

14. Bertozzi AL, Franco E, Mohler G, Short MB, Sledge D. 2020 The challenges of modeling and forecasting the spread of COVID-19. Proc. Natl. Acad. Sci. 117, 16732 LP –16738. (doi:10.1073/pnas.2006520117)

15. Holmdahl I, Buckee C. 2020 Wrong but useful — What Covid-19 epidemiologic models can and cannot tell us. N. Engl. J. Med. 383, 303–305. (doi:10.1056/NEJMp2016822)

16. Wood SN, Wit EC, Fasiolo M, Green PJ. 2021 COVID-19 and the difficulty of inferring epidemiological parameters from clinical data. Lancet Infect. Dis. 21, 27–28. (doi:10.1016/S1473-3099(20)30437-0)

17. Jalali MS, DiGennaro C, Sridhar D. 2020 Transparency assessment of COVID-19 models. Lancet Glob. Heal. 8, e1459–e1460. (doi:10.1016/S2214-109X(20)30447-2)

18. Smieszek T. 2009 A mechanistic model of infection: why duration and intensity of contacts should be included in models of disease spread. Theor. Biol. Med. Model. 6, 25. (doi:10.1186/1742-4682-6-25)

19. Drake JM et al. 2019 The statistics of epidemic transitions. PLOS Comput. Biol. 15, e1006917.

20. Scheffer M et al. 2009 Early-warning signals for critical transitions. Nature 461, 53–59. (doi:10.1038/nature08227)

21. Kéfi S, Dakos V, Scheffer M, Van Nes EH, Rietkerk M. 2013 Early warning signals also precede non-catastrophic transitions. Oikos 122, 641–648. (doi:https://doi.org/10.1111/j.1600-0706.2012.20838.x)

22. O’Regan SM, Drake JM. 2013 Theory of early warning signals of disease emergence and leading indicators of elimination. Theor. Ecol. 6, 333–357. (doi:10.1007/s12080-013-0185-5)

23. Carpenter SR et al. 2011 Early warnings of regime shifts: A whole-ecosystem experiment. Science (80-.). 332, 1079. (doi:10.1126/science.1203672)

24. Clements CF, Ozgul A. 2016 Including trait-based early warning signals helps predict population collapse. Nat. Commun. 7, 10984. (doi:10.1038/ncomms10984)

25. Brett T, Ajelli M, Liu Q-H, Krauland MG, Grefenstette JJ, van Panhuis WG, Vespignani A, Drake JM, Rohani P. 2020 Detecting critical slowing down in high-dimensional epidemiological systems. PLOS Comput. Biol. 16, e1007679.

26. Blumberg S, Lloyd-Smith JO. 2013 Inference of R0 and transmission heterogeneity from the size distribution of stuttering chains. PLOS Comput. Biol. 9, e1002993.

27. Miller PB, O’Dea EB, Rohani P, Drake JM. 2017 Forecasting infectious disease emergence subject to seasonal forcing. Theor. Biol. Med. Model. 14, 17. (doi:10.1186/s12976-017-0063-8)

28. Dibble CJ, O’Dea EB, Park AW, Drake JM. 2016 Waiting time to infectious disease emergence. J. R. Soc. Interface 13, 20160540. (doi:10.1098/rsif.2016.0540)

29. Harris MJ, Hay SI, Drake JM. 2020 Early warning signals of malaria resurgence in Kericho, Kenya. Biol. Lett. 16, 20190713. (doi:10.1098/rsbl.2019.0713)

30. Southall E, Tildesley MJ, Dyson L. 2020 Prospects for detecting early warning signals in discrete event sequence data: Application to epidemiological incidence data. PLOS Comput. Biol. 16, e1007836.

31. Kaur T, Sarkar S, Chowdhury S, Sinha SK, Jolly MK, Dutta PS. 2020 Anticipating the novel coronavirus disease (COVID-19) pandemic. Front. Public Heal.8, 521.

32. Pedro SA, Ndjomatchoua FT, Jentsch P, Tchuenche JM, Anand M, Bauch CT. 2020 Conditions for a second wave of COVID-19 due to interactions between disease dynamics and social processes. Front. Phys. 8, 428.

33. R Core Team. 2020 R: A language and environment for statistical computing.

34. Wood SN. 2011 Fast stable restricted maximum likelihood and marginal likelihood estimation of semiparametric generalized linear models. J. R. Stat. Soc. Ser. B (Statistical Methodol. 73, 3–36. (doi:https://doi.org/10.1111/j.1467-9868.2010.00749.x)

35. Burthe SJ et al. 2016 Do early warning indicators consistently predict nonlinear change in long-term ecological data? J. Appl. Ecol. 53, 666–676. (doi:https://doi.org/10.1111/1365-2664.12519)

36. McAloon C et al. 2020 Incubation period of COVID-19: a rapid systematic review and meta-analysis of observational research. BMJ Open 10, e039652. (doi:10.1136/bmjopen-2020-039652)

37. Simpson GL. 2021 gratia: Graceful ‘ggplot’-based graphics and other functions for GAMs fitted using ‘mgcv’.

38. Drake JM, Griffen BD. 2010 Early warning signals of extinction in deteriorating environments. Nature 467, 456–459. (doi:10.1038/nature09389)

39. Clements CF, Blanchard JL, Nash KL, Hindell MA, Ozgul A. 2017 Body size shifts and early warning signals precede the historic collapse of whale stocks. Nat. Ecol. Evol. 1, 188. (doi:10.1038/s41559-017-0188)

40. Dakos V et al.2012 Methods for detecting early warnings of critical transitions in time series illustrated using simulated ecological data. PLoS One 7, e41010. (doi:10.1371/journal.pone.0041010)

41. Dakos V, van Nes EH, D’Odorico P, Scheffer M. 2012 Robustness of variance and autocorrelation as indicators of critical slowing down. Ecology 93, 264–271. (doi:https://doi.org/10.1890/11-0889.1)

42. Wissel C. 1984 A universal law of the characteristic return time near thresholds. Oecologia 65, 101–107. (doi:10.1007/BF00384470)

43. Clements CF, McCarthy MA, Blanchard JL. 2019 Early warning signals of recovery in complex systems. Nat. Commun. 10, 1681. (doi:10.1038/s41467-019-09684-y)

44. O’Regan SM, Lillie JW, Drake JM. 2016 Leading indicators of mosquito-borne disease elimination. Theor. Ecol. 9, 269–286. (doi:10.1007/s12080-015-0285-5)

45. Clements CF, Drake JM, Griffiths JI, Ozgul A. 2015 Factors influencing the detectability of early warning signals of population collapse. Am. Nat. 186, 50–58. (doi:10.1086/681573)

46. Brett TS, O’Dea EB, Marty É, Miller PB, Park AW, Drake JM, Rohani P. 2018 Anticipating epidemic transitions with imperfect data. PLOS Comput. Biol. 14, e1006204.

47. Coccia M. 2021 The relation between length of lockdown, numbers of infected people and deaths of Covid-19, and economic growth of countries: Lessons learned to cope with future pandemics similar to Covid-19 and to constrain the deterioration of economic system. Sci. Total Environ. 775, 145801. (doi:https://doi.org/10.1016/j.scitotenv.2021.145801)

48. Dakos V, Carpenter SR, van Nes EH, Scheffer M. 2015 Resilience indicators: prospects and limitations for early warnings of regime shifts. Philos. Trans. R. Soc. B Biol. Sci. 370, 20130263. (doi:10.1098/rstb.2013.0263)

49. Wauchope HS, Amano T, Sutherland WJ, Johnston A. 2019 When can we trust population trends? A method for quantifying the effects of sampling interval and duration. Methods Ecol. Evol. 10, 2067–2078. (doi:https://doi.org/10.1111/2041-210X.13302)

