## Supplementary Information for "Early warning signals predict emergence of COVID-19 waves"

Supplementary Material: Early warning signals predict emergence of COVID-19 waves

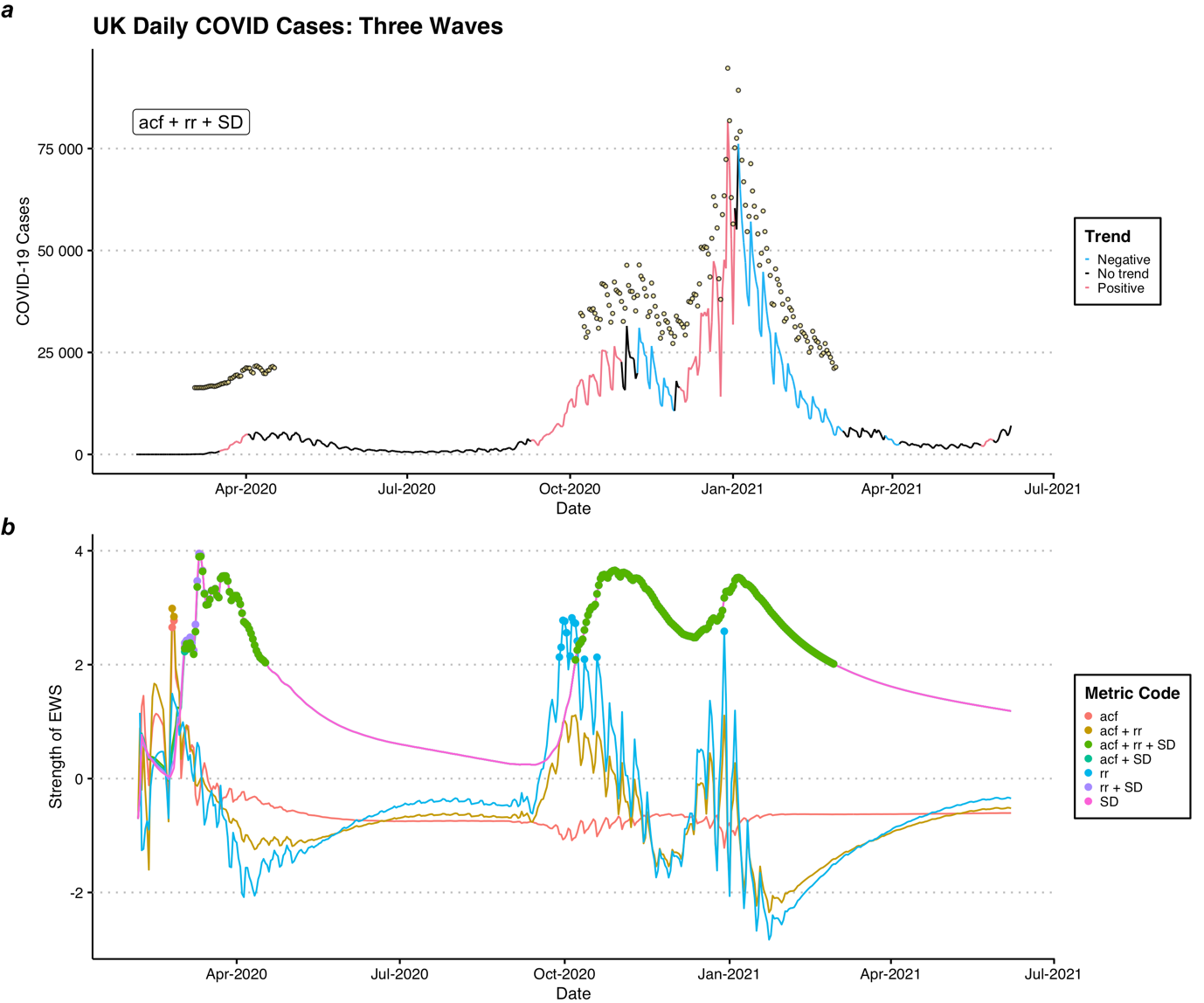

Supplementary Figure 1. Line plot of a) the daily UK COVD-19 cases and GAMM predicted trends, where yellow points represent detected early warning signals (EWS) from the triple composite indicator ‘acf + rr + SD’ experiencing continuous analysis, and b) individual EWS indicator strengths, where coloured dots indicate time points exceeding the 2σ threshold. No restarting of the EWS analysis was performed.

Supplementary Table 1. Summary of the first appearance and prediction of early warning indicators of UK COVID-19 case data under a) lenient and b) conservative definitions of ‘warning’. Reported indicators include: autocorrelation (acf), return rate (rr) and variance (SD).

| a) Early warning signal detection relative to wave onset. Single signal | | | | | | |
| --- | --- | --- | --- | --- | --- | --- |
| **Metric Code** | **Wave** | **Time of Wave Onset (Year-Month-Day)** | **Time of First EWS Detection (Year-Month-Day)** | **Number of Days Prior Signal Observed** | **Country** | **Prediction** |
| **acf** | first | 2020-03-17 | 2020-02-25 | 21 | UK | prior |
| **acf** | second | 2020-09-09 | 2020-08-06 | 34 | UK | prior |
| **acf** | third? | NA | 2021-06-04 | NA | UK | unknown |
| **acf + rr** | first | 2020-03-17 | 2020-02-25 | 21 | UK | prior |
| **acf + rr** | second | 2020-09-09 | 2020-07-20 | 51 | UK | prior |
| **acf + rr** | third? | NA | 2021-06-04 | NA | UK | unknown |
| **acf + rr + SD** | first | 2020-03-17 | 2020-03-03 | 14 | UK | prior |
| **acf + rr + SD** | second | 2020-09-09 | 2020-08-11 | 29 | UK | prior |
| **acf + rr + SD** | third? | NA | 2021-06-01 | NA | UK | unknown |
| **acf + SD** | first | 2020-03-17 | 2020-03-03 | 14 | UK | prior |
| **acf + SD** | second | 2020-09-09 | 2020-08-11 | 29 | UK | prior |
| **acf + SD** | third? | NA | 2021-06-01 | NA | UK | unknown |
| **rr** | first | 2020-03-17 | NA | NA | UK | missed |
| **rr** | second | 2020-09-09 | 2020-07-20 | 51 | UK | prior |
| **rr** | third? | NA | NA | NA | UK | match |
| **rr + SD** | first | 2020-03-17 | 2020-03-03 | 14 | UK | prior |
| **rr + SD** | second | 2020-09-09 | 2020-08-11 | 29 | UK | prior |
| **rr + SD** | third? | NA | 2021-06-01 | NA | UK | unknown |
| **SD** | first | 2020-03-17 | 2020-03-03 | 14 | UK | prior |
| **SD** | second | 2020-09-09 | 2020-08-11 | 29 | UK | prior |
| **SD** | third? | NA | 2021-06-01 | NA | UK | unknown |
| b) Early warning signal detection relative to wave onset. Consecutive signals | | | | | | |
| **Metric Code** | **Wave** | **Date Wave Onset (Year-Month-Day)** | **Date of First EWS Detection (Year-Month-Day)** | **Number of Days Prior Signal Observed** | **Country** | **Prediction** |
| **acf** | first | 2020-03-17 | NA | NA | UK | missed |
| **acf** | second | 2020-09-09 | NA | NA | UK | missed |
| **acf** | third? | NA | NA | NA | UK | match |
| **acf + rr** | first | 2020-03-17 | NA | NA | UK | missed |
| **acf + rr** | second | 2020-09-09 | NA | NA | UK | missed |
| **acf + rr** | third? | NA | NA | NA | UK | match |
| **acf + rr + SD** | first | 2020-03-17 | 2020-03-03 | 14 | UK | prior |
| **acf + rr + SD** | second | 2020-09-09 | 2020-08-11 | 29 | UK | prior |
| **acf + rr + SD** | third? | NA | 2021-06-01 | NA | UK | unknown |
| **acf + SD** | first | 2020-03-17 | 2020-03-03 | 14 | UK | prior |
| **acf + SD** | second | 2020-09-09 | 2020-08-11 | 29 | UK | prior |
| **acf + SD** | third? | NA | 2021-06-01 | NA | UK | unknown |
| **rr** | first | 2020-03-17 | NA | NA | UK | missed |
| **rr** | second | 2020-09-09 | NA | NA | UK | missed |
| **rr** | third? | NA | NA | NA | UK | match |
| **rr + SD** | first | 2020-03-17 | 2020-03-03 | 14 | UK | prior |
| **rr + SD** | second | 2020-09-09 | 2020-08-11 | 29 | UK | prior |
| **rr + SD** | third? | NA | 2021-06-01 | NA | UK | unknown |
| **SD** | first | 2020-03-17 | 2020-03-03 | 14 | UK | prior |
| **SD** | second | 2020-09-09 | 2020-08-11 | 29 | UK | prior |
| **SD** | third? | NA | 2021-06-01 | NA | UK | unknown |

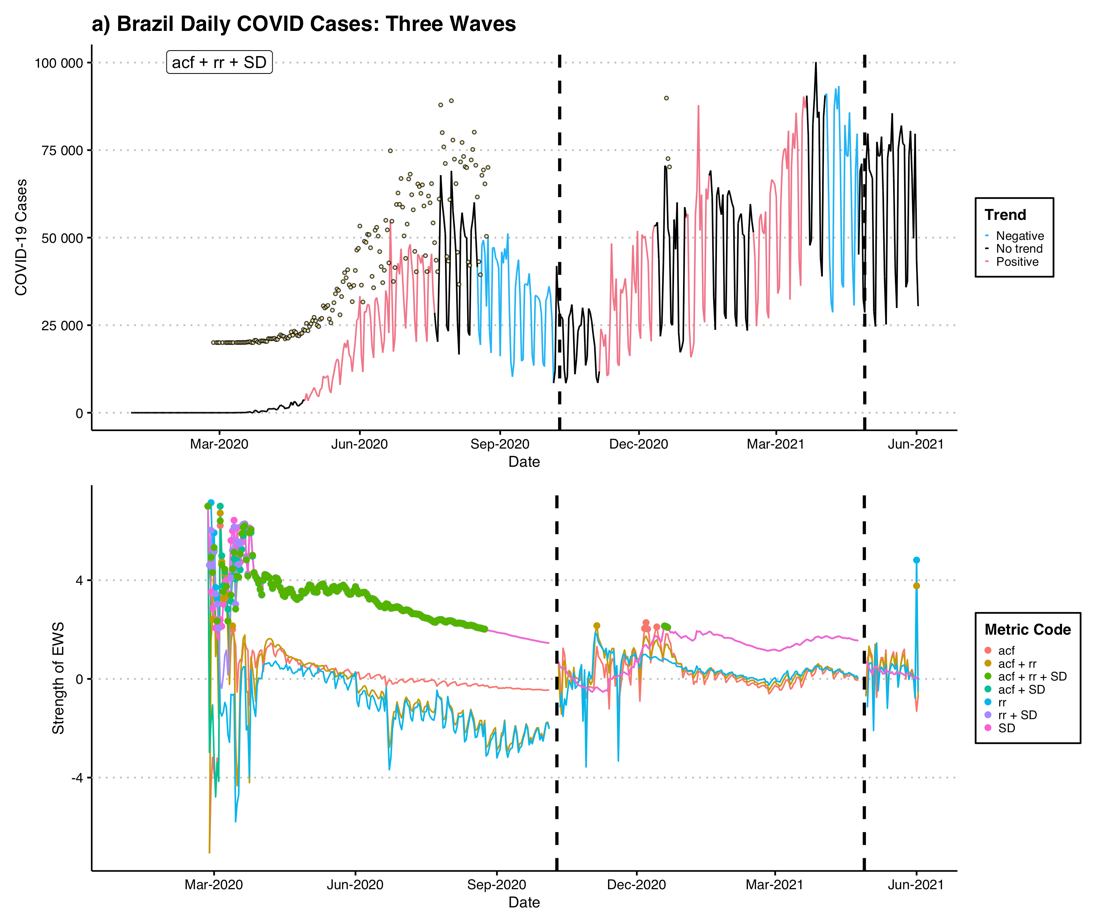

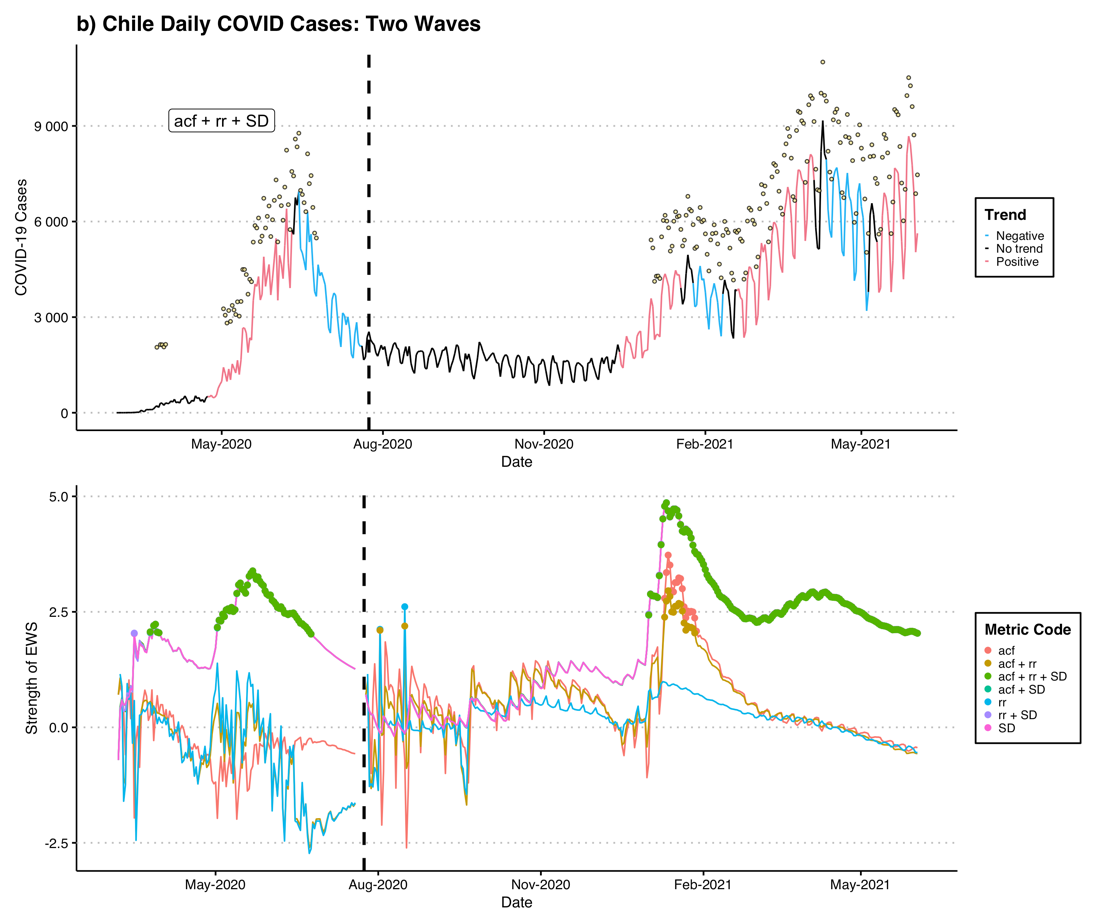

Supplementary Figure 2. Line plot of top) the daily country COVD-19 cases and GAMM predicted trends, where yellow points represent detected early warning signals (EWS) from the triple composite indicator ‘acf + rr + SD’, and bottom) individual EWS indicator strengths, where coloured dots indicate time points exceeding the 2σ threshold. Dashed, vertical lines indicate time points of EWS reassessment.

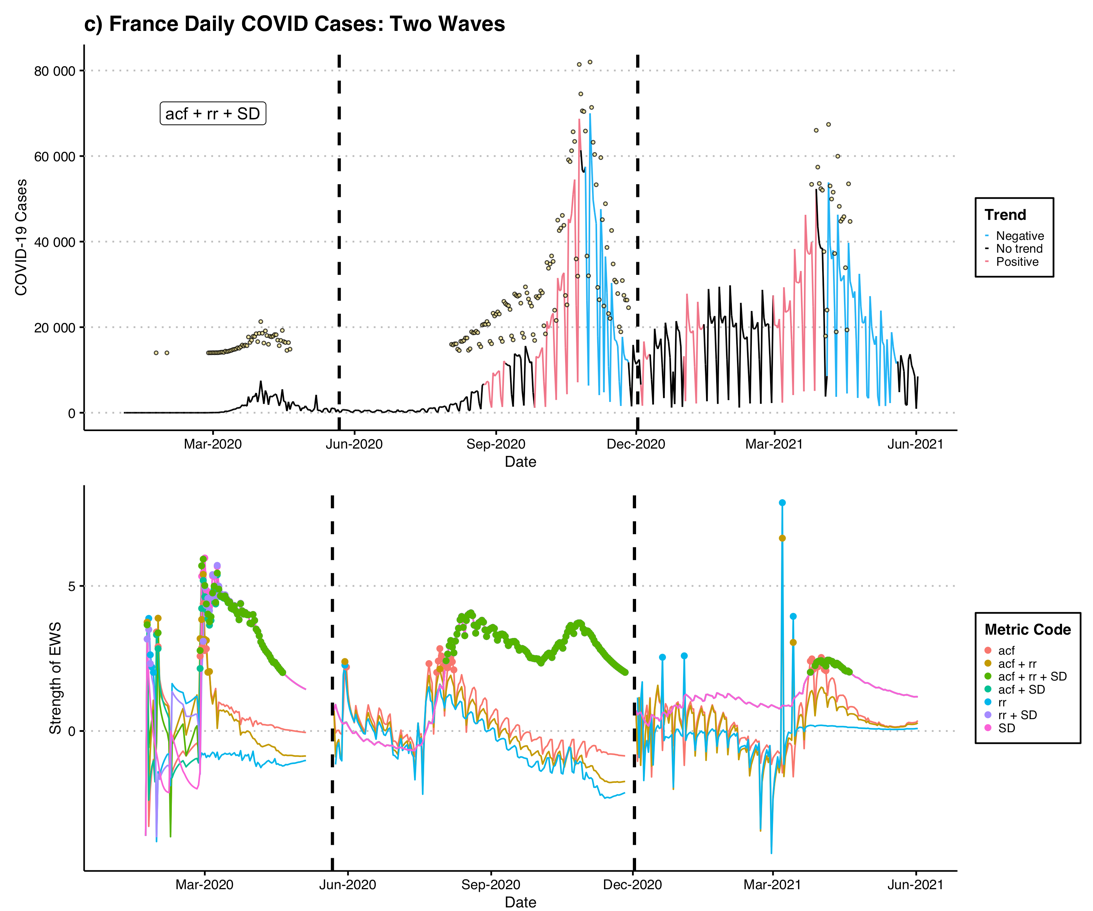

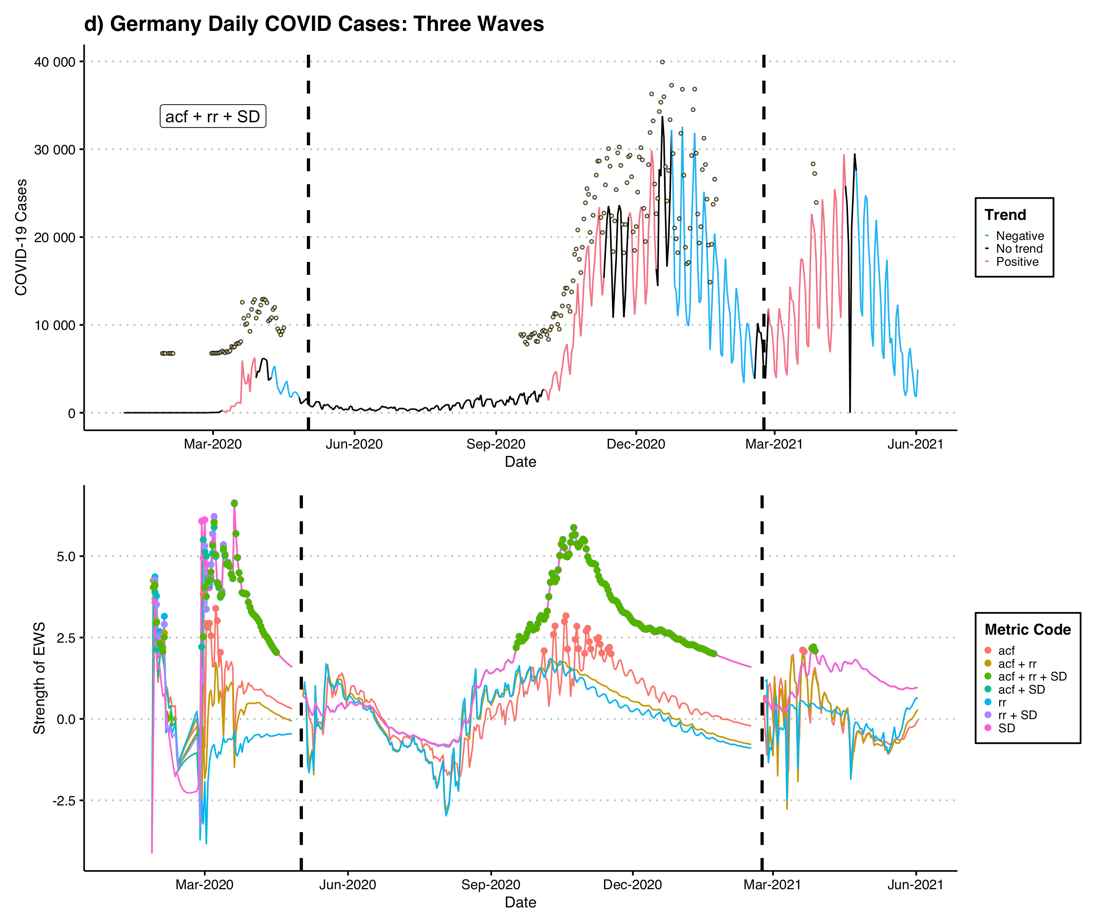

Supplementary Figure 2 cont.

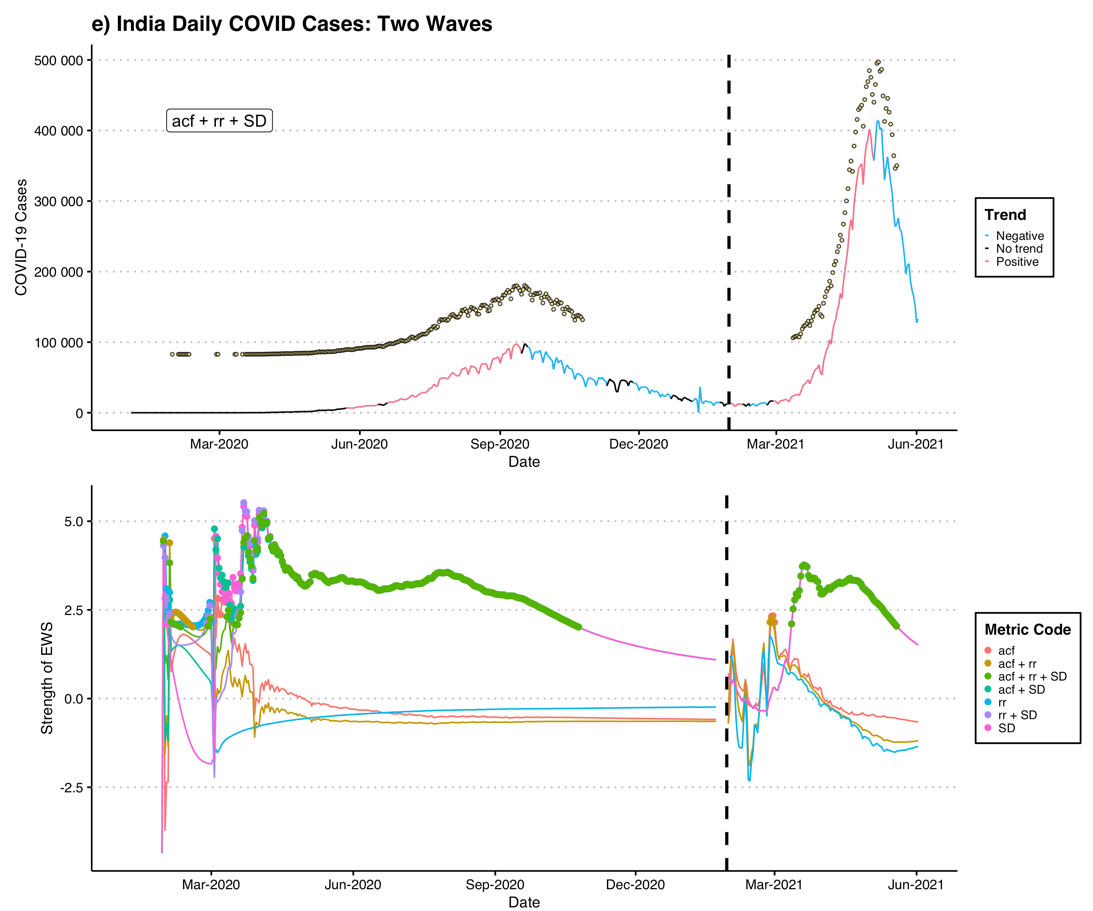

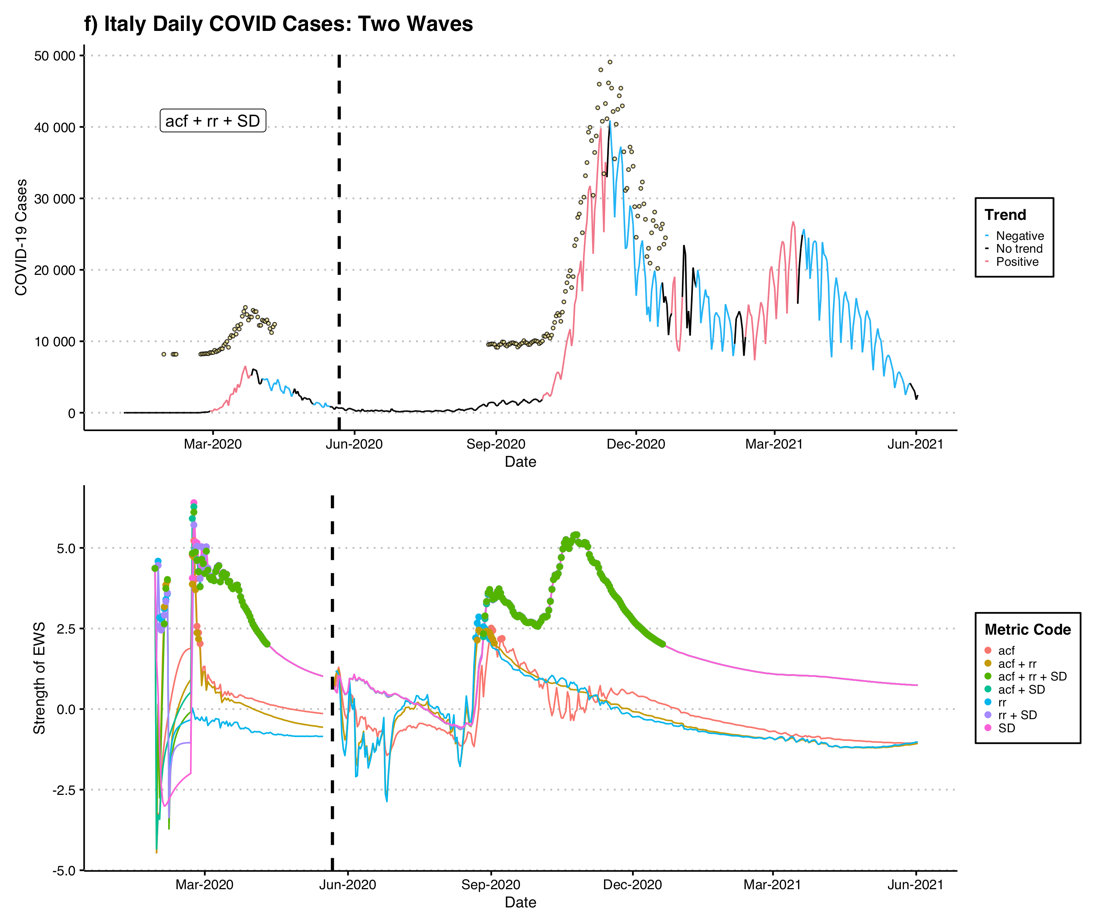

Supplementary Figure 2 cont.

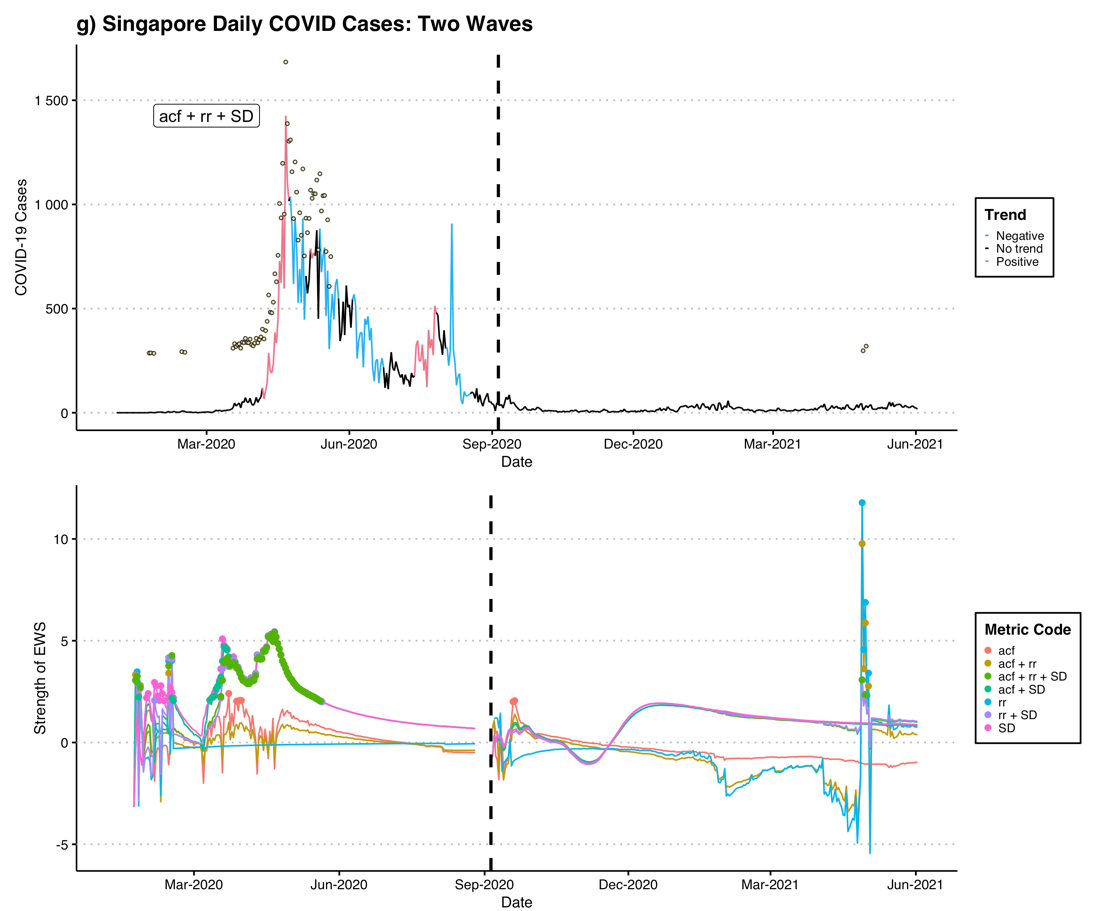

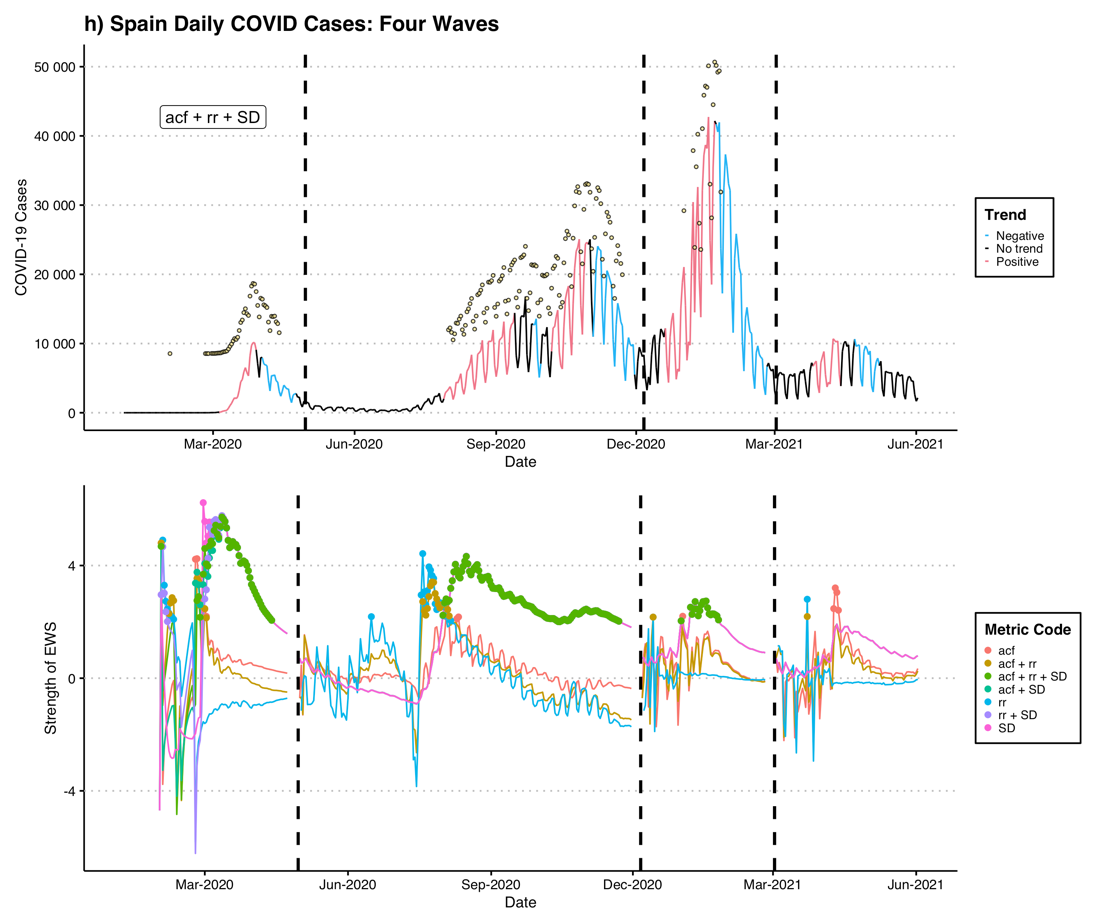

Supplementary Figure 2 cont.

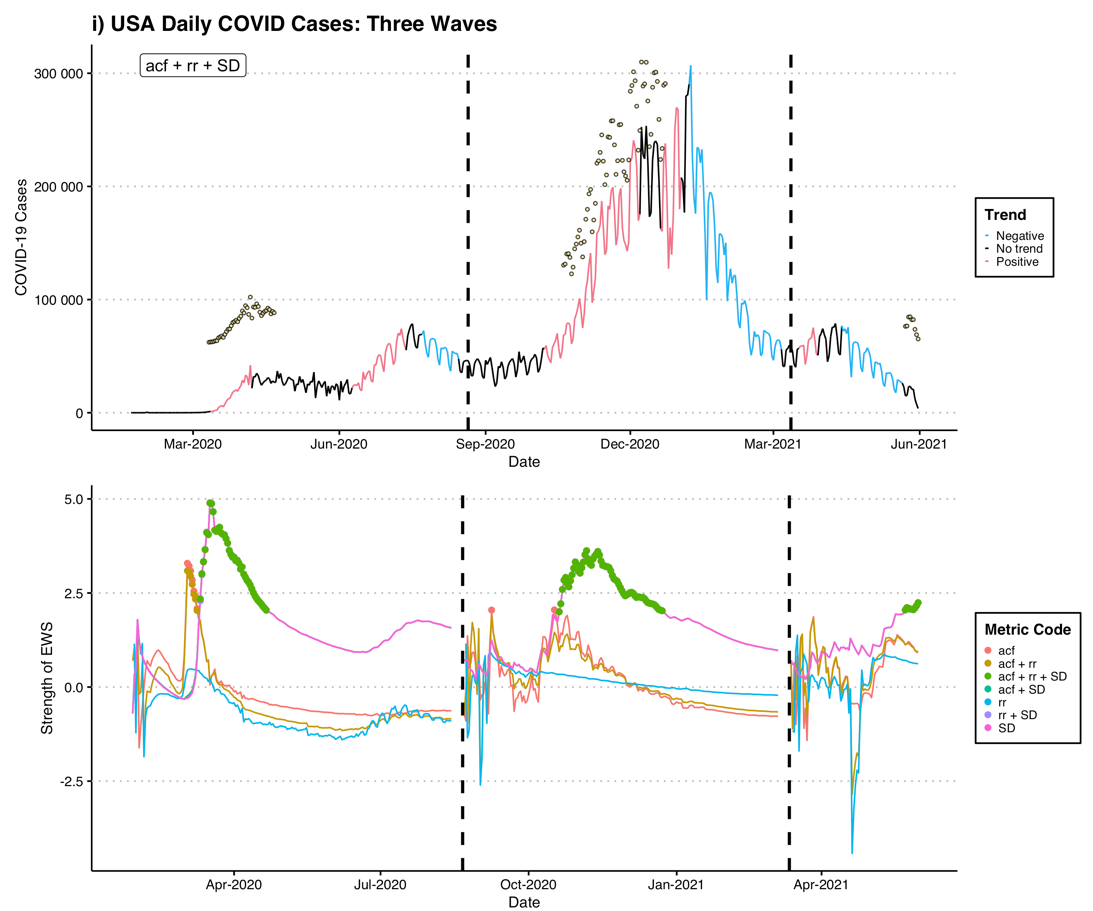

Supplementary Figure 2 cont.
